# *HHBayes*: A Flexible Bayesian Framework for Simulating and Analyzing Household Transmission Dynamics

**DOI:** 10.64898/2026.04.01.26349903

**Authors:** Ke Li, Yiren Hou, Bhramar Mukherjee, Virginia E. Pitzer, Daniel M. Weinberger

**Author notes:** These authors contributed equally to this work (Ke Li, Yiren Hou).

## Abstract

Household transmission studies are important for understanding infectious disease transmission and evaluating interventions; however, they are frequently constrained by methodological challenges, including in study design and sample size determination, and in estimating parameters of interest after collecting the data. Existing tools often lack flexibility in modeling age-specific susceptibility, infectivity patterns, and the impact of interventions such as vaccination or prophylaxis. Here, we develop *HHBayes*, an open-source R package that provides a unified framework for simulating and analyzing household transmission data using Bayesian methods. The package enables researchers to: (1) simulate realistic household transmission dynamics with highly customizable variables; (2) incorporate viral load data (measured in viral copies/mL or cycle threshold values) to model time-varying infectiousness; (3) estimate age-dependent susceptibility and infectivity parameters using Hamiltonian Monte Carlo methods implemented in Stan; and (4) evaluate intervention effects through user-defined covariates that modify susceptibility or infectivity. We demonstrate the capabilities of the package through simulation studies showing accurate parameter recovery and applications to seasonal respiratory virus transmission, including the impact of vaccination and antiviral prophylaxis on household attack rates. *HHBayes* addresses a critical gap in infectious disease epidemiology by providing researchers with accessible tools for both prospective study design and retrospective data analysis. The flexibility of the package in handling complex household structures, time-varying infectiousness, and intervention effects makes it valuable for studying diverse pathogens.

**Author Summary:** Understanding how infections spread within households is important to controlling infectious diseases, yet designing and analyzing household transmission studies remains challenging. We developed *HHBayes*, a comprehensive R package that helps researchers both plan new studies and analyze existing data. The package can simulate realistic household transmission data with different family structures, incorporate the effects of vaccines or treatments, and account for how infectiousness changes over the course of infection using viral load data. Researchers can then use Bayesian statistical methods to estimate how susceptible different age groups are to infection and how infectious they become when infected. We demonstrate that the package successfully recovers known parameters in simulations and can handle real-world complexities like reinfections and seasonal patterns. This tool will be particularly valuable for studying respiratory viruses like influenza and respiratory syncytial virus, where understanding household transmission is crucial for evaluating intervention strategies.

## Introduction

Households are key settings for infectious disease transmission [1]. Understanding transmission dynamics within households is essential for multiple public health objectives, including estimating the basic reproduction number and secondary attack rates (SAR), and predicting and evaluating the effectiveness of vaccines and interventions [2,3]. For respiratory pathogens such as influenza, SARS-CoV-2, and respiratory syncytial virus (RSV), household studies have provided invaluable insights into age-specific attack rates, transmission heterogeneity, and the protection conferred by prior immunity [4–9].

Despite their epidemiological importance, household transmission studies pose methodological challenges. First, transmission events are rarely observed directly; instead, researchers must infer who infected whom from serial testing data and symptom onset [10,11]. Second, heterogeneity exists in both susceptibility (the probability of becoming infected upon exposure) and infectivity (the probability of transmitting infection to others), which varies by age, contact patterns, immune status, pathogen characteristics, and interventions [11,12]. The variability complicates parameter identifiability and increases uncertainty in transmission inference and effect estimation.

Several methodological frameworks exist for analyzing household transmission data, each with specific strengths and limitations. Chain binomial models provide simple, analytically tractable approaches for estimating secondary attack rates, but assume homogeneous susceptibility and infectivity and do not account for multiple generations of within-household transmission, which can introduce bias in SAR estimates [3,13,14]. Bayesian data-augmentation approaches address these limitations by imputing unobserved infection times within an MCMC framework, enabling estimation of age-specific susceptibility and infectiousness while accounting for incomplete observation of the transmission process [10,15]. Building on this foundation, transmission hazard models with regression adjustment further allow simultaneous estimation of within-household and external forces of infection alongside individual-level risk factors [16]. More sophisticated statistical approaches, such as those implemented in *outbreaker2* [17] and *epicontacts [18]*, enable transmission chain reconstruction from temporal and contact data but typically lack integration with viral load information and do not provide unified frameworks for simulation and inference.

No existing tools provide an integrated framework for both simulating household transmission studies during the design phase and analyzing real data with Bayesian inference. This gap leaves researchers without standardized methods for power calculations, parameter estimation under realistic household structures, or evaluation of intervention scenarios. Furthermore, the increasing availability of high-resolution viral kinetics data from quantitative polymerase chain reaction (qPCR) testing (cycle threshold (Ct) values) or viral load quantification remains underutilized in household transmission models, despite its potential to improve transmission inference [19–21].

Here, we developed *HHBayes* to address these methodological gaps through a comprehensive, flexible, and user-friendly R package. The package is designed to be accessible to researchers with basic R programming experience while providing advanced users the flexibility to customize household structures, viral kinetics models, and prior distributions. By integrating simulation and inference in a single framework, *HHBayes* enables iterative study design, power analysis, and hypothesis generation, followed by rigorous analysis of collected data. The package provides four core functionalities:

1. **Flexible simulation framework:** Generates realistic household transmission data with user-specified household structures (e.g., single vs. two-parent families, presence of grandparents, number of siblings), seasonal forcing patterns, testing frequency, and other pathogen-specific parameters, such as infectious periods. The simulation engine implements a stochastic SIRS-like model with age-specific susceptibility and infectivity parameters, viral load trajectories based on gamma distributions or empirical Ct value curves, and household-level contact matrices.
2. **Intervention modeling:** Flexibly incorporates covariates that modify susceptibility, infectivity, or both. This enables researchers to simulate and evaluate the impact of vaccines (reducing susceptibility), antiviral treatments (reducing infectivity), prophylaxis, behavioral interventions, or combinations. Coverage levels can be specified by age group, allowing realistic modeling of age-targeted interventions.
3. **Bayesian parameter estimation:** Estimates age-dependent susceptibility and infectivity parameters, baseline transmission rates, and covariate effects in a Bayesian framework. The package handles complex data structures including multiple infections per individual and time-varying viral load observations.
4. **Visualization and inference tools:** Generates epidemic curves, transmission network diagrams at the household level, posterior distributions of estimated parameters, and visualizations for reconstructed transmission chains within households. Built-in functions support model validation by enabling direct comparisons between simulated and fitted data, as well as systematic assessment of parameter recovery.

## Methods

### Mathematical Model for Household Transmission

The *HHBayes* package implements a discrete-time stochastic transmission model that incorporates both within-household and community transmission pathways. We model individuals as transitioning between susceptible (*S*), infectious (*I*), and recovered/immune (*R*) states, with explicit tracking of infection times, viral load trajectories, and reinfection events after waning immunity. Recovered individuals become susceptible after immunity wanes.

### Force of Infection

For individual *i* in household *h* at time *t*, the instantaneous hazard of infection (force of infection, λ) is given by:

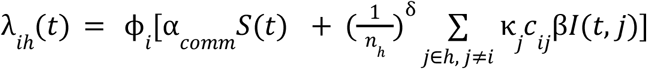

where:

- ϕ_*i*_ is the susceptibility parameter for individual *i*,
- α_*comm*_ is the baseline community transmission rate,
- *S*(*t*) is a seasonal forcing term,
- *n*_*h*_ is the number of individuals in household *h* (*n*_*h*_ ≥ 1),
- δ is a scaling factor for the impact of household size on the household hazard,
- κ _*j*_ is the infectivity parameter for infectious household member *j*,
- *c* _*ij*_ is the contact matrix between individuals *i* and *j*,
- β*I*(*t, j*) is the infectiousness from an infected individual *j* at time *t*, and β*I*(*t, j*) = β_1_ + β_2_ *f*(·) where:
  ∘ β_1_ represents baseline transmission rate during the infection period
  ∘ β_2_ *f*(·) represents additional transmission modulated by:
    ▪ *f*(*V*_*j*_ (*t*)): viral load-dependent infectiousness when viral load data are available
    ▪ *f* (*t* − *t* _*j*_): a gamma distribution describing infectiousness since infection when viral load data are not available.

### Viral Load Integration

The package supports two approaches for modeling time-varying infectiousness. For viral load data measured as log10-scaled copies/mL, we model individual trajectories using a biphasic model:

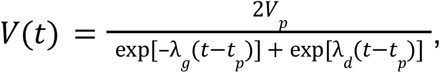

where *V*_*p*_ is peak viral load, *t*_*p*_ is time to peak, λ_*g*_ is growth rate, λ_*d*_ is decay rate, and *t* is time since infection. These parameters can vary by different factors, such as age group. The default viral load parameter values are taken from a previous modeling study [20]. The contribution to infectiousness is:

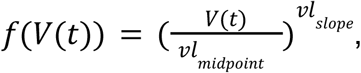

Where *vl*_*midpoint*_ is the reference log10-scaled viral load values for infectiousness, and *vl*_*midpoint*_ is power-law exponent for log10-scaled viral load scaling. For viral load data measured as Ct values, we model trajectories as:

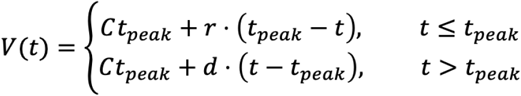

where *Ct*_*peak*_ is the minimum Ct value (maximum viral load), *r* is the rate of decline before peak, *d* is the rate of increase after peak, and *t*_*peak*_ is the day of peak viral load. The contribution to infectiousness is:

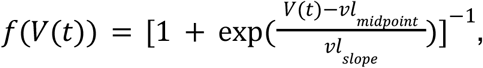

where *vl*_*midpoint*_ here is the Ct value at which infectiousness is 50% of its maximum, and *vl*_*slope*_ determines the steepness of the curve. We choose this functional form because it provides a good fit to RSV viral load data [20].

### Covariate Effects

To model interventions or individual-level characteristics, the package implements covariate modifiers on susceptibility and infectivity:

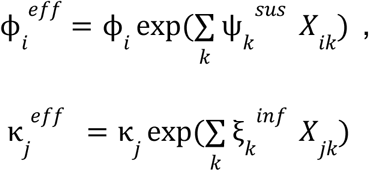

where *X*_*ik*_ is a matrix of *k* covariates for individual *i* (e.g., vaccination status), ψ_*k*_ ^*sus*^ and ξ_*k*_ ^*inf*^ are coefficients representing log-scale effects to modify susceptibility and infectivity, respectively. This parameterization allows interventions to proportionally reduce susceptibility or infectivity.

### Infection Process and State Transitions

At each time step, susceptible individuals become infected with probability:

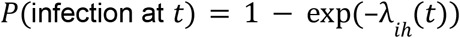

Upon infection, individuals transition to the infectious state and are assigned a specific viral load trajectory that belongs to group *i*. The infectious period is drawn from a gamma distribution. Following recovery, individuals acquire temporary immunity with a gamma-distributed duration, after which immunity wanes and they return to the susceptible state, allowing for reinfection.

### Simulation Algorithm

**simulate_multiple_households_comm()**, which integrates household generation, infection dynamics, and diagnostic sampling within a unified simulation framework. For each household over the study period, all individuals are initialized as susceptible unless specified otherwise, and susceptible individuals experience daily risk of infection from both community transmission and infectious household members, with household exposure weighted by viral load and contact patterns. Infection events occur stochastically based on the combined force of infection. Newly infected individuals are assigned viral load trajectories drawn from age-specific parameter distributions, remain infectious for durations sampled from user-defined gamma distributions, and subsequently have temporary immunity with waning times also drawn from user-defined gamma distributions. Once immunity wanes, individuals return to the susceptible state, enabling reinfection.

The simulation incorporates flexible household composition generated stochastically using the **generate_household_roles()** function based on user-specified demographic profiles. The simulation function also allows users to define flexible surveillance and testing schemes, including scheduled PCR testing, symptom-triggered testing following the first detected case within a household, and customizable test characteristics such as viral load or Ct thresholds for positivity, with options for perfect or imperfect detection. Other variables, including the contact matrix between household members and covariates such as vaccination status, can also be defined in the function.

### Bayesian Inference Framework

Parameter estimation is performed using Hamiltonian Monte Carlo (HMC) method via the Stan probabilistic programming language, accessed through the **prepare_stan_data()** function and a custom Stan model. For each susceptible individual *i* in household *h* observed from time *t*_*start*_ to infection time *t*_*inf*_ (or end of study *T*), the likelihood contribution is:

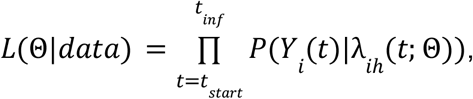

Where *Y*_*i*_ (*t*) = 1 if individual *i* is infected at time *t*, or 0 otherwise, and the estimated parameters are Θ = {ϕ_*i*_, κ_*j*_, β_1_, β_2_, α, β^*cov*^, *vl*_*midpoint*_, *vl*_*slope*_}. When viral load data are not in use, the estimated parameters are Θ = {ϕ_*i*_, κ_*j*_, β_1_, β_2_, α, β ^*cov*^, *gen*_*shape*_, *gen*_*rate*_}, where *gen*_*shape*_ and *gen*_*rate*_ are parameters for a gamma distribution describing infectiousness since infection. The package supports flexible prior specification, allowing model parameters to be estimated under normal, uniform, or lognormal distributions. The model is fitted using HMC with the No-U-Turn Sampler, running four independent chains by default. Each chain includes 1,000 warm-up iterations followed by 1,000 post-warm-up sampling iterations. Convergence is assessed using standard diagnostics, with satisfactory performance defined by 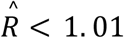 (Gelman-Rubin diagnostic) and effective sample sizes exceeding 1,000, ensuring adequate mixing and sampling efficiency. Posterior samples enable full uncertainty quantification for all parameters, including derived quantities like secondary attack rates and intervention effectiveness.

Computational time depends on data size and model complexity. For a typical dataset of 50 households without covariates, model fitting using four HMC chains (1,000 warm-up and 1,000 sampling iterations per chain) requires approximately 12 minutes on a laptop equipped with a 10-core Apple M1 Pro CPU and 16 GB RAM. The computational time increases to approximately 20 minutes for a dataset of 100 households. Parallel execution of chains across cores substantially reduces wall-clock time.

### Data Preparation and Imputation

Real household transmission data often lack precise information on infection and recovery times. To address this, the **prepare_stan_data()** function applies a stochastic imputation procedure to infer individual infection intervals. For each infection episode, the infection start day is sampled uniformly between the date of the most recent negative test and the date of the first positive test, while the infection end day is sampled uniformly between the date of the last positive test and the date of the subsequent negative test. Based on these imputed interval-censored data, the function constructs individual-specific risk windows that identify periods during which each person is at risk of infection. This approach accommodates longitudinal household data with potentially multiple infection episodes per individual while remaining computationally tractable. See https://github.com/keli5734/HHBayes for detailed variable and covariate specification.

### Visualization and Diagnostic Tools

The package includes a comprehensive set of visualization and summary tools to support model evaluation and interpretation. The **plot_epidemic_curve()** function generates a dual-axis plot that compares simulated infections, colored by age group, with observed surveillance data aggregated over user-specified time intervals. Posterior uncertainty is summarized using **plot_posterior_distributions()**, which produces violin plots of posterior samples for the susceptibility and infectivity parameters to facilitate comparisons across age groups. Covariate effects are displayed with **plot_covariate_effects()** as forest plots with 95% credible intervals. Transmission dynamics at the household level are illustrated using **plot_household_timeline()**, a visualization function to reconstruct household transmission chains. In addition, **summarize_attack_rates()** computes primary attack rates and reinfection rates stratified by age group. Together, these tools support both model validation in simulation studies and interpretation of results from real-world data analyses.

## Results

### Simulation Study 1: Parameter Recovery and Model Validation

We first validated the package’s ability to recover known parameters through a simulation-estimation study. We simulated 100 households over a 365-day period. Household composition was generated according to the following distribution: every household included one infant and two parents; the number of additional siblings was assigned with probabilities of 0, 1, or 2 siblings at 0%, 80%, and 20%, respectively; and the number of grandparents present followed probabilities of 0, 1, or 2 at 70%, 10%, and 20%, respectively. We assumed a one-year study period with simulated surveillance data representing the force of infection from the community. We assumed that infants and toddlers are three times as susceptible as adults, while they are two times as infectious as adults (i.e., the default reference group in the function). Sampling was performed at four-day intervals. We ran the following code to generate the simulated dataset and obtain summary statistics for the attack rates.

~~~
*<preformat>*
  *household_profile <- list(prob_infant = 1.0*,
  *prob_adults = c(0, 0, 1)*,
  *prob_siblings = c(0, .8, .2)*,
  *prob_elderly = c(0.7, 0.1, 0.2))*
*study_start <- “2024-07-01”*
*study_end <- “2025-06-30”*
*dates_weekly <- seq(from = as.Date(study_start), to = as.Date(study_end), by = “week”)*
*set.seed(123)*
*surveillance_data <- data.frame(*
  *date = dates_weekly*,
  *cases = 0.1 + 100 * exp(−0.0002 * (as.numeric(dates_weekly - mean(dates_weekly)))^2) +
abs(rnorm(length(dates_weekly),mean = 0, sd = 10)))*
*sim_res <- simulate_multiple_households_comm(*
  *n_households = 100*,
  *viral_testing = “viral load”*,
  *phi_by_role = c(adult = 1, infant = 3, toddler = 3, elderly = 1)*,
  *kappa_by_role = c(adult = 1, infant = 2, toddler = 2, elderly = 1)*,
  *infectious_shape = 7*,
  *infectious_scale = 1*,
  *waning_shape = 16*,
  *waning_scale = 10*,
  *surveillance_interval = 4*,
  *start_date = study_start*,
  *end_date = study_end*,
  *surveillance_df = surveillance_data*,
  *seed = 100*,
  *household_profile_list = household_profile)*
*rates <- summarize_attack_rates(sim_res)*
*plot_epidemic_curve(sim_res, surveillance_data, start_date_str = study_start, bin_width = 7)*
~~~

The main output of the simulation data has the following format (**Table 1**):

**Table 1.**
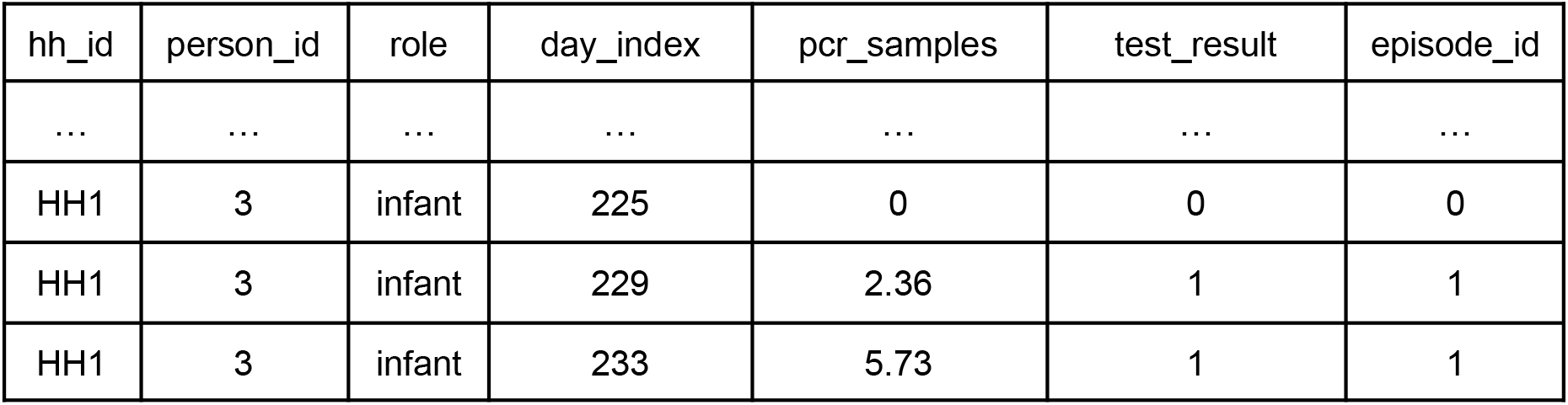
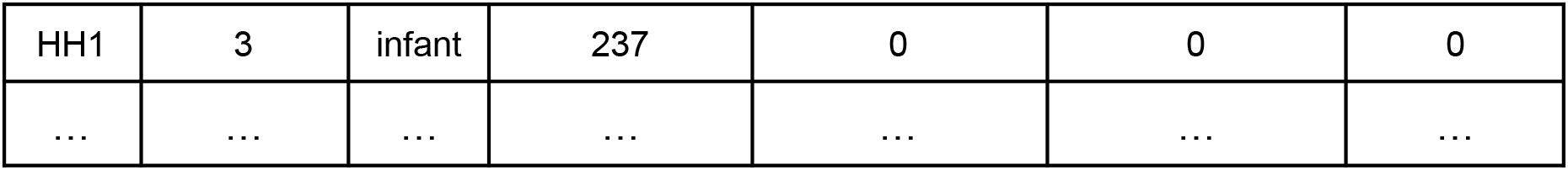
The format of simulated household transmission dataset.

| hh_id | person_id | role | day_index | pcr_samples | test_result | episode_id |
| --- | --- | --- | --- | --- | --- | --- |
| ... | ... | ... | ... | ... | ... | ... |
| HH1 | 3 | infant | 225 | 0 | 0 | 0 |
| HH1 | 3 | infant | 229 | 2.36 | 1 | 1 |
| HH1 | 3 | infant | 233 | 5.73 | 1 | 1 |
| HH1 | 3 | infant | 237 | 0 | 0 | 0 |
| ... | ... | ... | ... | ... | ... | ... |

The dataset includes a household ID (*hh_id*, categorical), a person ID (*person_id*, numeric), each individual’s role (*role*, categorical: infant, toddler, adult, elderly), the day relative to the study start (day_index, numeric), viral load measurements (*pcr_samples*, numeric; log10-scaled viral copies/ml or Ct values), test results (*test_result*, numeric; 0 = negative, 1 = positive), and an episode ID (*episode_id*, numeric; 0 = not infected, 1 = first episode, 2 = second episode). If users have a real dataset, the package expects it to be organized in this format.

We compared the temporal dynamics of the incidence in the study cohort with simulated surveillance data using the function **plot_epidemic_curve()** (**Fig. 1**). We found the overall attack rate is 45% (219 out of 487). Most infections occurred among infants (attack rate (AR), 51% (51/100)) and toddlers (attack rate, 55.1% (70/127)). Attack rates were lower among adults at 37% (74/200) and older adults at 40% (24/60). We also observed two reinfection episodes among toddlers.

**Figure 1.**
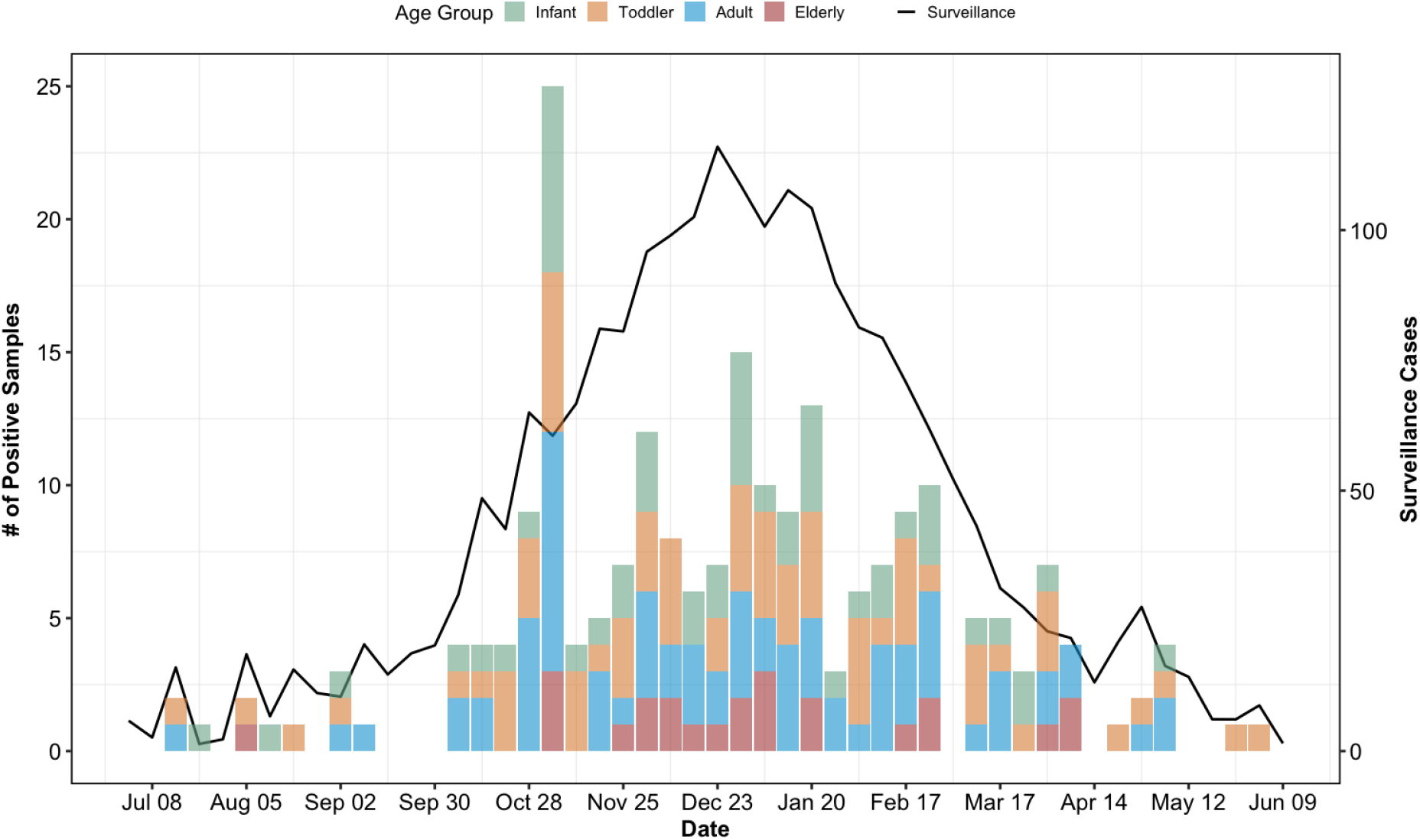
Temporal distribution of simulated positive samples in the study cohort compared to simulated community surveillance data. The stacked bar chart (left y-axis) shows the daily count of positive samples collected within the study, stratified by age group: Infant (green), Toddler (orange), Adult (blue), and Elderly (red). The solid grey line (right y-axis) represents the weekly number of detections from community surveillance.

We then fitted the model to our simulated dataset by running the code:

~~~
*df_for_stan <- sim_res$diagnostic_df*
*my_priors <- list(*
  *beta1 = list(dist = “normal”, params = c(−5, 1))*,
  *beta2 = list(dist = “normal”, params = c(−5, 1))*,
  *alpha = list(dist = “normal”, params = c(−7, 1))*,
  *phi_role = list(dist = “normal”, params = c(0, 0.5))*,
  *kappa_role = list(dist = “normal”, params = c(0, 0.5))*,
  *vl_midpoint = list(dist = “normal”, params = c(3, .2))*,
  *vl_slope = list(dist = “normal”, params = c(2.5, 0.2)))*
*stan_input <- prepare_stan_data(*
  *df_clean = df_for_stan*,
  *surveillance_df = surveillance_data*,
  *study_start_date = as.Date(study_start)*,
  *study_end_date = as.Date(study_end)*,
  *use_vl_data = TRUE*,
  *use_curve_logic = FALSE*,
  *delta = 0*,
  *priors = my_priors)*
*options(mc.cores = parallel::detectCores())*
*fit <- fit_household_model(stan_input)*
~~~

We found that the model successfully recovered all key parameters, with narrow credible intervals (**Fig. 2**). Age-specific susceptibility and infectivity parameters were estimated with high certainty for the infant and toddler groups. In contrast, estimates of susceptibility and infectivity for the older age group were relatively biased, likely reflecting the small number of infected older adults in the simulated dataset (n = 24). Susceptibility and infectivity parameters for adults were fixed at 1 and used as reference values.

**Figure 2.**
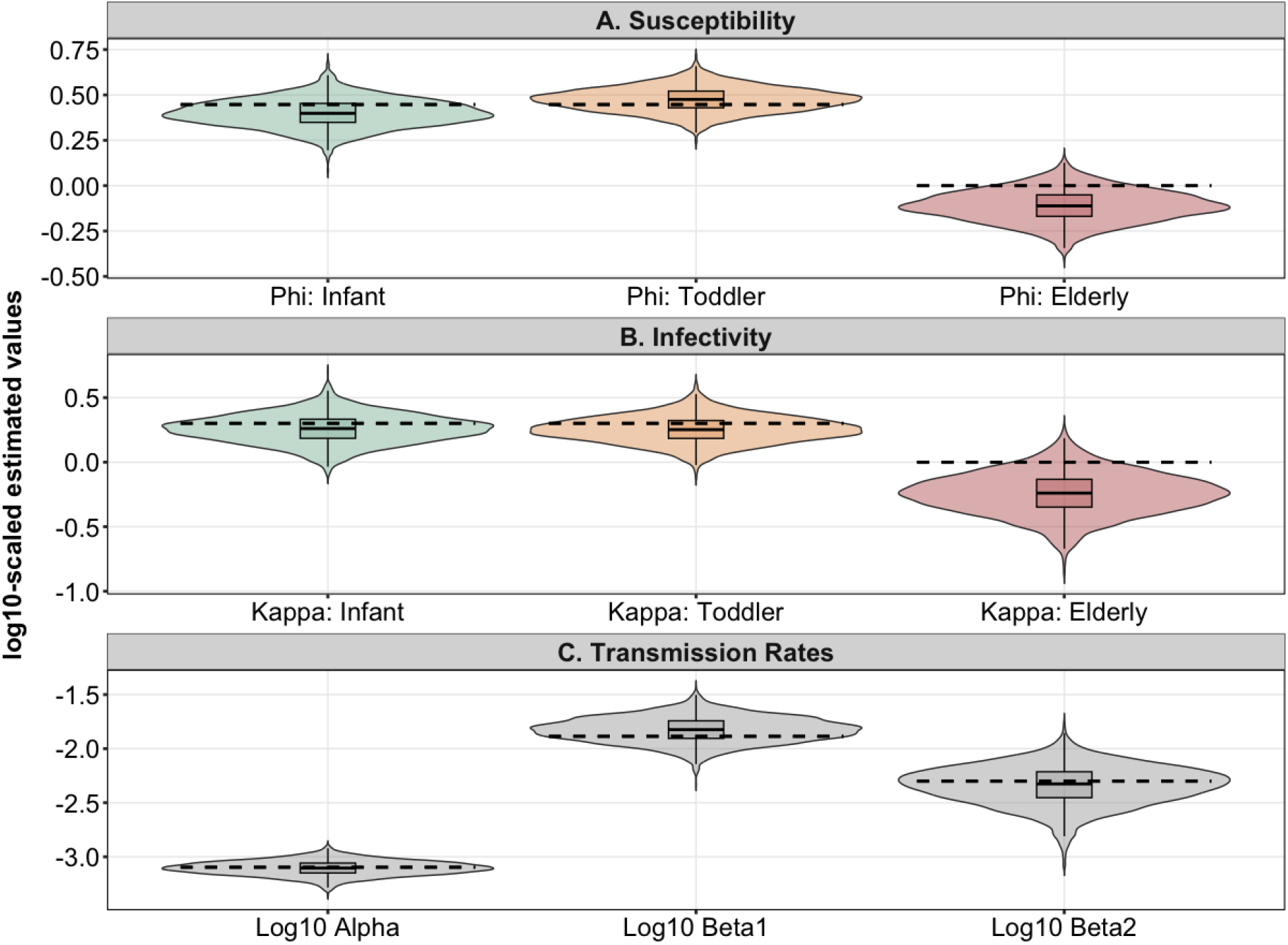
Age-specific transmission parameters and transmission rates. Posterior distributions of log10-scaled **(A)** relative susceptibility (ϕ) and **(B)** relative infectivity by role group (κ) and **(C)** transmission rates. Violin plots show the full density of the estimates, with internal boxplots indicating the median and interquartile range. A total of 4,000 samples were drawn after the burn-in period. Dashed lines indicate the true parameter values.

All analyses were conducted on a laptop with a 10-core Apple M1 Pro CPU and 16 GB RAM. Fitting the model with four chains required approximately 20 minutes for the simulated dataset analyzed here.

Using the estimated household transmission parameters, we can reconstruct household transmission chains by running the code:

~~~
*# 2. Reconstruct Chains*
*chains <- reconstruct_transmission_chains(fit, stan_input)*
*# 3. Plot a specific Household (e.g*., *HH #5)*
*p_hh <- plot_household_timeline(chains, stan_input, target_hh_id = 5)*
*print(p_hh)*
~~~

Household transmission chains allow us to reconstruct both the order and timing of infections within households (**Fig. 3**). For example in Household 5, the transmission dynamics reveal a sequential outbreak starting in mid February. The initial chain begins when a toddler acts as the index case, acquiring the infection directly from the community (100% probability). This introduction drives the spread within the home: the toddler likely transmitted the virus to both the infant and adult with 98.2% and 52.7% certainty, respectively. The probability the adult acquired infection from the infant was 46.8%.

**Figure 3.**
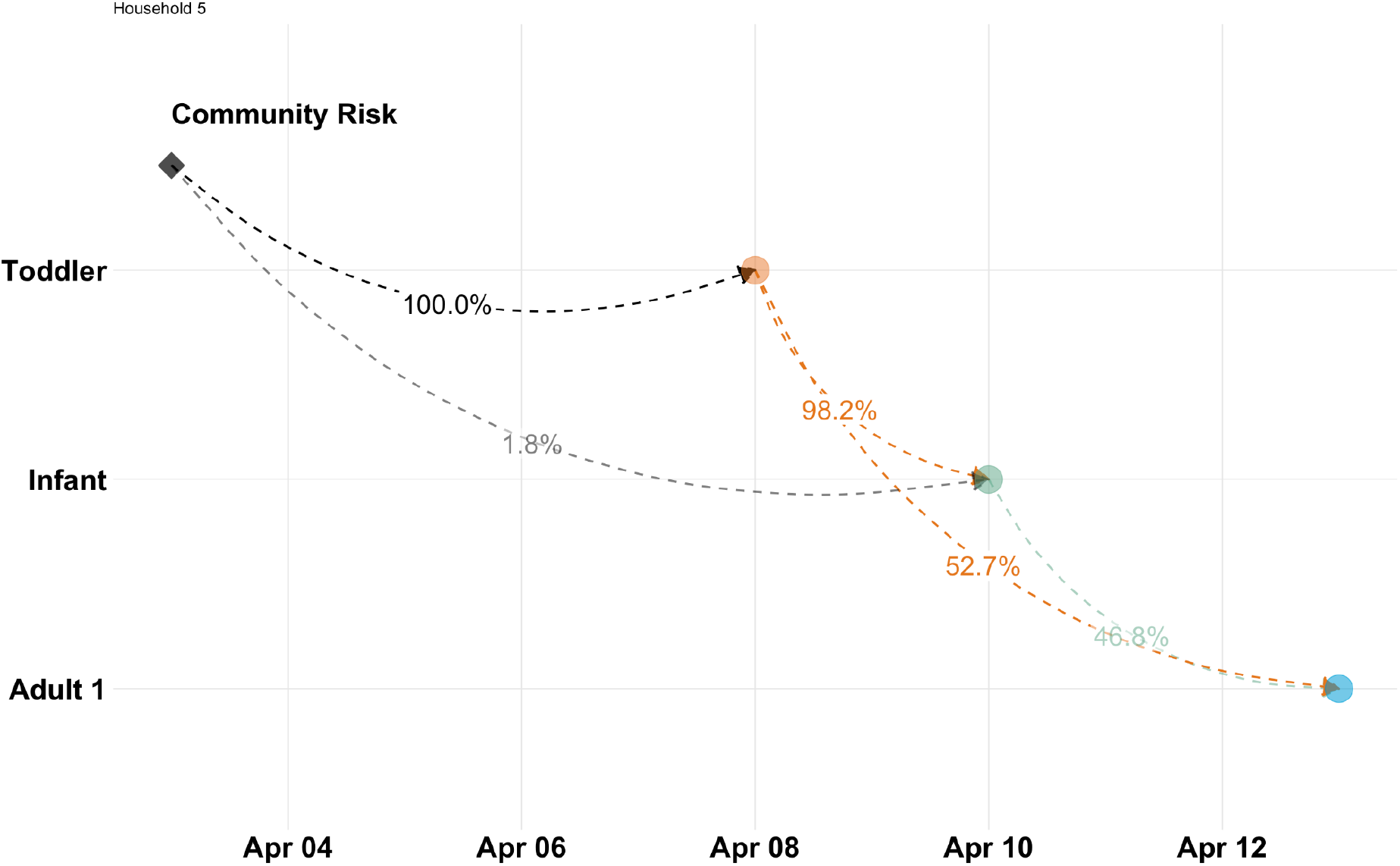
Reconstructed transmission pathways within a selected household. The vertical axis represents individual household members, while the horizontal axis indicates the estimated date of infection. Black diamonds denote the introduction of pathogens from the community (index cases). Colored circles represent infection events by roles. Dashed arrows indicate the direction of transmission between individuals; colors of the arrows are associated with the infector; the associated percentages reflect the inferred probability of that specific transmission event occurring. Faint grey lines represent lower-probability transmission sources.

### Simulation Study 2: Application to Intervention Modeling

We also demonstrate the covariate functionality of the package using a simulated household-randomized immunization intervention involving 200 households. The immunization intervention was assumed to reduce susceptibility to infection by 80% and reduce infectivity of infected individuals by 80%, with 80% coverage among infants only; all other age groups were assumed to remain unprotected. To simulate a dataset with the intervention and estimate the intervention effect, we defined the relevant covariates directly in the **prepare_stan_data()** function as follows:

~~~
*sim_config <- list(*
  *list(name=“vacc_status”*,
     *efficacy=.8*,
     *effect_on=“both”*,
     *coverage=list(infant=.8, toddler=0, adult=0, elderly=0)))*
*sim_res <- simulate_multiple_households_comm(*
     *n_households = 200*,
     *surveillance_interval = 4*,
     *start_date = study_start*,
     *end_date = study_end*,
     *surveillance_df = surveillance_data*,
     *covariates_config = sim_config)*
*person_covariates <- sim_res$hh_df %>%*
     *dplyr::select(hh_id, person_id, vacc_status) %>% dplyr::distinct()*
*df_for_stan <- sim_res$diagnostic_df %>%*
     *dplyr::left_join(person_covariates, by = c(“hh_id”, “person_id”))*
 *my_priors <- list(*
 *beta1 = list(dist = “normal”, params = c(−5, 1))*,
 *beta2 = list(dist = “normal”, params = c(−5, 1))*,
 *alpha = list(dist = “normal”, params = c(−7, 1))*,
 *phi_role = list(dist = “normal”, params = c(0, 1))*,
 *kappa_role = list(dist = “normal”, params = c(0, 1))*,
 *vl_midpoint = list(dist = “normal”, params = c(3*, . *2))*,
 *vl_slope = list(dist = “normal”, params = c(2.5, 0.2)))*
*stan_input <- prepare_stan_data(*
 *df_clean = df_for_stan*,
 *surveillance_df = surveillance_data*,
 *study_start_date = as.Date(study_start)*,
 *study_end_date = as.Date(study_end)*,
 *covariates_susceptibility = c(“vacc_status”)*,
 *covariates_infectivity = c(“vacc_status”)*,
 *use_vl_data = TRUE*,
 *use_curve_logic = FALSE*,
 *delta = 0*,
 *priors = my_priors)*
*fit <- fit_household_model(stan_input)*
~~~

We found that the model successfully recovered the effects of the immunization intervention on both susceptibility and infectivity (**Fig. 4**). The posterior median of the covariate coefficient for reduced susceptibility was −1.82 (95% credible interval (CI): [−2.61, −1.04]), corresponding to an estimated effectiveness of 83.8% (95% CI: [64.7%, 92.6%]) in reducing susceptibility to infection. The posterior median of the coefficient of the intervention effect on infectivity was −1.51 (95% CI: [−2.45, 0.52]) yielding an estimated effectiveness of 77.8% (95% CI: [−68.2%, 91.4%]). These insignificant estimates likely reflect the limited information available to identify infectivity effects, which rely on observing secondary transmission events in the households and are therefore harder to estimate precisely given the sample size and household structure, particularly because immunization was assumed to cover infants only.

**Figure 4.**
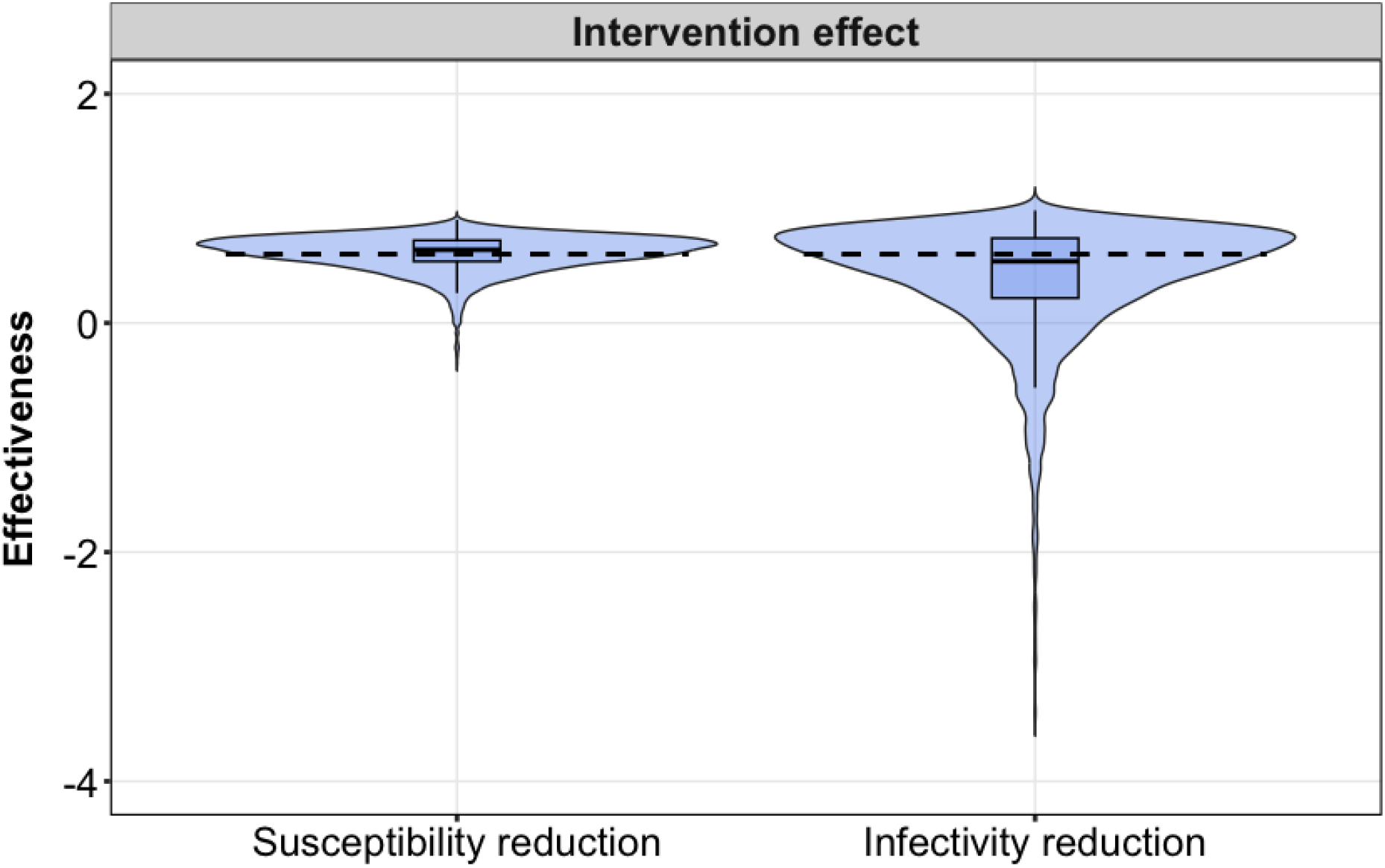
Estimates of intervention effects. Posterior distributions of intervention effects to reduce susceptibility to infection and infectivity. Violin plots show the full density of the estimates, with internal boxplots indicating the median and interquartile range. A total of 4,000 samples were drawn after the burn-in period. Dashed lines indicate the true parameter values.

## Discussion

Here, we developed and demonstrated *HHBayes*, a comprehensive R package for simulating and analyzing household transmission data within a Bayesian framework. Through a simulation–estimation study, we demonstrated accurate parameter recovery and illustrated how intervention strategies can be incorporated into the model and estimated from the simulated data. The package addresses critical gaps in infectious disease epidemiology by providing researchers with accessible tools for both prospective study design and retrospective data analysis, with particular strengths in incorporating age heterogeneity, viral kinetics data, and potential interventions into household transmission studies.

*HHBayes* supports both prospective and retrospective household transmission studies, providing tailored tools for each design. Prospective study designs facilitate the collection of data on asymptomatic transmission from active monitoring, while retrospective approaches allow for the rapid analysis of existing transmission data. For prospective study designs, our package provides an integrated simulation–inference framework that allows researchers to design studies, perform power calculations, and analyze data within a single environment, ensuring consistency between design assumptions and analytical approaches. The package supports incorporation of viral kinetics data by explicitly modeling time-varying infectiousness based on viral load measured by viral copies/mL or Ct value trajectories, improving the accuracy of transmission inference, particularly in settings with frequent sampling [20–23]. It also features a flexible covariate system in which user-defined covariates can modify susceptibility, infectivity, or both, enabling evaluation of diverse interventions, such as vaccines and antivirals, without modifying the core model code. In addition, *HHBayes* enables direct definition and estimation of age-group-specific susceptibility and infectivity parameters, which are often of primary epidemiological interest but challenging to identify from household data due to confounding between age, contact patterns, and household size [11,24–26]. The package also allows users to tailor simulations to targeted demographic settings by specifying household structures and contact probabilities between individuals. Assuming uniform family structures and homogeneous contact patterns may underestimate the sample size required to detect age-specific effects and may lead to biased parameter estimates [11]. Finally, the package supports reinfection modeling through explicit representation of immunity waning and reinfection dynamics, an increasingly important feature for pathogens with short-lived immunity, such as SARS-CoV-2 and RSV [27,28]. For retrospective studies, the package can analyze existing household infection data, reconstruct likely transmission chains, and estimate age-specific susceptibility and infectivity even when sampling is incomplete or irregular [10].

Given the unique features of the package, we believe *HHBayes* is particularly well-suited for a variety of research contexts. First, in study design and power analysis, researchers planning household transmission studies can simulate various scenarios to determine the sample sizes needed to detect specific effects [29]. For example, the package could be used to estimate the number of participants required to detect a 40% vaccine efficacy with 80% power by simulating data under both the null and alternative hypotheses and examining confidence intervals. Second, for testing interventions, the package can be used to estimate both direct and indirect effects in household-randomized trials of vaccines, antivirals, or behavior changes [30]. Its flexible covariate system works for situations where only some household members get the intervention, as well as when entire households are assigned. Third, for hypothesis generation, simulating transmission under different mechanistic assumptions, such as age-dependent versus age-independent infectivity or short versus long immunity duration, can produce predictions that guide data collection and inform model selection in subsequent analyses [31,32].

The package has some limitations. First, the package implements a discrete-time SIRS-like model with gamma-distributed infectious and immunity periods. While these assumptions are reasonable for many respiratory pathogens, they may not capture all biological complexity. For instance, the model does not explicitly include latent periods, which may be important for pathogens with substantial pre-infectious shedding. Future versions could incorporate more flexible models. Second, computational demands may limit scalability for very large studies or highly parameterized models, as the current HMC-based inference can become slow when handling thousands of households, multiple age groups, or numerous covariates.

In conclusion, *HHBayes* provides an advanced statistical toolkit for infectious disease epidemiology, providing researchers with accessible, flexible, and rigorous methods for studying pathogen transmission in the household setting. By unifying simulation and inference in a Bayesian framework, the package enables researchers to design better studies, estimate age-specific transmission parameters, evaluate intervention effects, and generate hypotheses about transmission mechanisms.

## Data Availability

All code for HHBayes is available under an open-source MIT license at https://github.com/keli5734/HHBayes. The package includes extensive documentation, vignettes demonstrating common workflows, and example datasets. We encourage researchers to contribute extensions, report bugs, and share use cases through the GitHub issue tracker.

## Acknowledgments

We thank the developers of Stan, RStan, and the tidyverse packages for creating the software infrastructure that makes *HHBayes* possible.

## Supporting Information

### S1 Code: Reproducible Analysis Scripts

Annotated R scripts reproducing all figures in the manuscript.

## Code Availability

All code for *HHBayes* is available under an open-source MIT license at https://github.com/keli5734/HHBayes. The package includes extensive documentation, vignettes demonstrating common workflows, and example datasets. We encourage researchers to contribute extensions, report bugs, and share use cases through the GitHub issue tracker.

## References

1. Møgelmose S, Vijnck L, Neven F, Neels K, Beutels P, Hens N. Population age and household structures shape transmission dynamics of emerging infectious diseases: a longitudinal microsimulation approach. J R Soc Interface. 2023;20: 20230087. doi:10.1098/rsif.2023.0087

2. Wang C, Huang X, Lau EHY, Cowling BJ, Tsang TK. Association between population-level factors and household secondary attack rate of SARS-CoV-2: A systematic review and meta-analysis. Open Forum Infect Dis. 2023;10: ofac676. doi:10.1093/ofid/ofac676

3. Prunas O, Warren JL, Crawford FW, Gazit S, Patalon T, Weinberger DM, et al. Vaccination with BNT162b2 reduces transmission of SARS-CoV-2 to household contacts in Israel. Science. 2022;375: 1151–1154. doi:10.1126/science.abl4292

4. Funk A, Florin TA, Kuppermann N, Finkelstein Y, Kazakoff A, Baldovsky M, et al. Household transmission dynamics of asymptomatic SARS-CoV-2-infected children: A multinational, controlled case-ascertained prospective study. Clin Infect Dis. 2024;78: 1522–1530. doi:10.1093/cid/ciae069

5. Cohen C, McMorrow ML, Martinson NA, Kahn K, Treurnicht FK, Moyes J, et al. Cohort profile: A Prospective Household cohort study of Influenza, Respiratory syncytial virus and other respiratory pathogens community burden and Transmission dynamics in South Africa, 2016-2018. Influenza Other Respi Viruses. 2021;15: 789–803. doi:10.1111/irv.12881

6. Kombe IK, Munywoki PK, Baguelin M, Nokes DJ, Medley GF. Model-based estimates of transmission of respiratory syncytial virus within households. Epidemics. 2019;27: 1–11. doi:10.1016/j.epidem.2018.12.001

7. Munywoki PK, Koech DC, Agoti CN, Lewa C, Cane PA, Medley GF, et al. The source of respiratory syncytial virus infection in infants: A household cohort study in rural Kenya. J Infect Dis. 2014;209: 1685–1692. doi:10.1093/infdis/jit828

8. Munywoki PK, Koech DC, Agoti CN, Kibirige N, Kipkoech J, Cane PA, et al. Influence of age, severity of infection, and co-infection on the duration of respiratory syncytial virus (RSV) shedding. Epidemiol Infect. 2015;143: 804–812. doi:10.1017/S0950268814001393

9. Hall CB, Geiman JM, Biggar R, Kotok DI, Hogan PM, Douglas GR Jr. Respiratory syncytial virus infections within families. N Engl J Med. 1976;294: 414–419. doi:10.1056/NEJM197602192940803

10. Cauchemez S, Carrat F, Viboud C, Valleron AJ, Boëlle PY. A Bayesian MCMC approach to study transmission of influenza: application to household longitudinal data. Stat Med. 2004;23: 3469–3487. doi:10.1002/sim.1912

11. Layan M, Hens N, de Hoog MLA, Bruijning-Verhagen PCJL, Cowling BJ, Cauchemez S. Addressing current limitations of household transmission studies by collecting contact data. Am J Epidemiol. 2024;193: 1832–1839. doi:10.1093/aje/kwae106

12. Boldea O, Alipoor A, Pei S, Shaman J, Rozhnova G. Age-specific transmission dynamics of SARS-CoV-2 during the first 2 years of the pandemic. PNAS Nexus. 2024;3: pgae024. doi:10.1093/pnasnexus/pgae024

13. Longini IM Jr, Koopman JS, Monto AS, Fox JP. Estimating household and community transmission parameters for influenza. Am J Epidemiol. 1982;115: 736–751. doi:10.1093/oxfordjournals.aje.a113356

14. Sharker Y, Kenah E. Estimating and interpreting secondary attack risk: Binomial considered biased. PLoS Comput Biol. 2021;17: e1008601. doi:10.1371/journal.pcbi.1008601

15. Lau MSY, Cowling BJ, Cook AR, Riley S. Inferring influenza dynamics and control in households. Proc Natl Acad Sci U S A. 2015;112: 9094–9099. doi:10.1073/pnas.1423339112

16. Crawford FW, Marx FM, Zelner J, Cohen T. Transmission modeling with regression adjustment for analyzing household-based studies of infectious disease: Application to tuberculosis. Epidemiology. 2020;31: 238–247. doi:10.1097/EDE.0000000000001143

17. Jombart T, Cori A, Didelot X, Cauchemez S, Fraser C, Ferguson N. Bayesian reconstruction of disease outbreaks by combining epidemiologic and genomic data. PLoS Comput Biol. 2014;10: e1003457. doi:10.1371/journal.pcbi.1003457

18. Nagraj VP, Randhawa N, Campbell F, Crellen T, Sudre B, Jombart T. epicontacts: Handling, visualisation and analysis of epidemiological contacts. F1000Res. 2018;7: 566. doi:10.12688/f1000research.14492.2

19. Hay JA, Kennedy-Shaffer L, Kanjilal S, Lennon NJ, Gabriel SB, Lipsitch M, et al. Estimating epidemiologic dynamics from cross-sectional viral load distributions. Science. 2021;373: eabh0635. doi:10.1126/science.abh0635

20. Li K, Bont LJ, Weinberger DM, Pitzer VE. Relating in vivo RSV infection kinetics to host infectiousness in different age groups. J Infect Dis. 2025; jiaf138. doi:10.1093/infdis/jiaf138

21. Ke R, Zitzmann C, Ho DD, Ribeiro RM, Perelson AS. In vivo kinetics of SARS-CoV-2 infection and its relationship with a person’s infectiousness. medRxiv. 2021. doi:10.1101/2021.06.26.21259581

22. Jones TC, Biele G, Mühlemann B, Veith T, Schneider J, Beheim-Schwarzbach J, et al. Estimating infectiousness throughout SARS-CoV-2 infection course. Science. 2021;373. doi:10.1126/science.abi5273

23. Marc A, Kerioui M, Blanquart F, Bertrand J, Mitjà O, Corbacho-Monné M, et al. Quantifying the relationship between SARS-CoV-2 viral load and infectiousness. Elife. 2021;10. doi:10.7554/eLife.69302

24. Davies NG, Klepac P, Liu Y, Prem K, Jit M, CMMID COVID-19 working group, et al. Age-dependent effects in the transmission and control of COVID-19 epidemics. Nat Med. 2020;26: 1205–1211. doi:10.1038/s41591-020-0962-9

25. Toth DJA, Sheets TR, Beams AB, Ahmed SM, Seegert N, Love J, et al. Model-based estimates of age-structured SARS-CoV-2 epidemiology in households. medRxiv. medRxiv; 2024. doi:10.1101/2024.04.18.24306047

26. Cauchemez S, Donnelly CA, Reed C, Ghani AC, Fraser C, Kent CK, et al. Household transmission of 2009 pandemic influenza A (H1N1) virus in the United States. N Engl J Med. 2009;361: 2619–2627. doi:10.1056/NEJMoa0905498

27. Goldman JD, Wang K, Röltgen K, Nielsen SCA, Roach JC, Naccache SN, et al. Reinfection with SARS-CoV-2 and waning humoral immunity: A case report. Vaccines (Basel). 2022;11: 5. doi:10.3390/vaccines11010005

28. Baker RE, Park SW, Yang W, Vecchi GA, Metcalf CJE, Grenfell BT. The impact of COVID-19 nonpharmaceutical interventions on the future dynamics of endemic infections. Proceedings of the National Academy of Sciences. 2020;117: 30547–30553. doi:10.1073/pnas.2013182117

29. Arnold BF, Hogan DR, Colford JM Jr, Hubbard AE. Simulation methods to estimate design power: an overview for applied research. BMC Med Res Methodol. 2011;11: 94. doi:10.1186/1471-2288-11-94

30. Cowling BJ, Chan K-H, Fang VJ, Cheng CKY, Fung ROP, Wai W, et al. Facemasks and hand hygiene to prevent influenza transmission in households: a cluster randomized trial. Ann Intern Med. 2009;151: 437–446. doi:10.7326/0003-4819-151-7-200910060-00142

31. Metcalf CJE, Lessler J. Opportunities and challenges in modeling emerging infectious diseases. Science. 2017;357: 149–152. doi:10.1126/science.aam8335

32. Cobey S. Modeling infectious disease dynamics. Science. 2020;368: 713–714. doi:10.1126/science.abb5659

